# Cardio Heart Connect: Protocol for a Randomized Trial of a Commercially Available mHealth Fitness Intervention for Cardiac Rehabilitation After Transcatheter Aortic Valve Replacement

**DOI:** 10.64898/2026.06.16.26355836

**Authors:** Joshua Rosen, Neel Butala, Daniel B. Kramer, Laura Helmkamp, Kristy Gama, Elizabeth Goldberg, Lucas Marzec, Pamela N Peterson, Madeline Balser, Duncan Riley, Sean Vongvivitpatana, Ilian Mino, Emily Greenway, Christopher E. Knoepke, Jennifer D. Portz

## Abstract

**Background:** Despite ample evidence of the benefits of cardiac rehabilitation (CR), few transcatheter aortic valve replacement (TAVR) patients participate. Commercially available mobile health offers an opportunity to deliver activity-promotion content to populations that are challenged to participate in CR. This study aims to test the efficacy of clinically controlled, commercially available fitness programming for improving physical activity and cardiovascular health outcomes designed to be initiated while patients are on waitlists for traditional CR.

**Methods:** The Cardio Heart Connect study is a hybrid type I effectiveness-implementation trial aiming to enroll N=200 patients who have been placed on a cardiac rehab waitlist following a TAVR procedure from the University of Colorado Hospital Heart and Vascular Center. Participants will be randomized 1:1 to the Cardio Heart Connect intervention with commercially available fitness or attention control, designed to control for technology access. At baseline, post-intervention (8 weeks), and follow-up (12 months), we will assess the primary outcome of participants’ daily steps as measured by smartwatch accelerometer and secondary outcomes of interest including functional capacity (Duke Activity Status Index; VO2max), quality of life (Kansas City Cardiomyopathy Questionnaire), and cardiovascular health status (Life Essential 8). In addition, we will use mixed methodologies to evaluate the implementation of intervention using the Reach, Effectiveness, Adoption, Implementation, and Maintenance (RE-AIM) Framework.

**Conclusions:** Commercially available fitness programs have the potential to provide more accessible opportunities for patients recovering from TAVR to engage in physical activity and may be preferred due to their customizability, convenience, and ease of scheduling. Overall, this study will provide insight into the use of commercial mHealth to promote activity following TAVR.

## Introduction

Cardiac rehabilitation (CR) is a Class 1A recommendation for hundreds of thousands of patients per year, including those who have recently undergone percutaneous valve procedures, based on evidence that participation improves mortality, functional status, and exercise capacity.^1^ Despite ample evidence of the benefits of CR only a fraction (∼29%)^2^ of patients receiving transcatheter aortic valve replacement (TAVR) participate, which is thought to be a combination of missed referral opportunities coupled with patient-centric treatment burdens.^3^ Moreover, many CR-eligible patients are placed on waitlists following referral, missing a critical window to improve functional capacity when motivation is high. Participation in CR is particularly low among older TAVR patients with comorbidities, women, racial/ethnic minorities, and those with lower SES or living in rural areas, contributing to disproportionate morbidity and mortality burden in these patients.^4^ These waitlists are a function of capacity; even if all CR programs in the US were operating above capacity, they could serve less than half of all eligible patients. While virtual and remote CR programs convey clinical improvements similar to facility-based programs,^5, 6, 7^ they are not yet widely available.

Commercially available fitness apps, including Peloton, Zwift, Strava, and others, can provide at-home opportunities for structured activity promotion for these patients. Moreover, patients may prefer them due to their customizability, convenience, and ease of scheduling. Remote monitoring of activity, safety, and physiological well-being is possible via smart watches, improving the viability of at-home exercise.^8–10^ These applications operate on the principle that content can be delivered to patients 1) immediately, 2) via technology they already use, 3) at a time and place that is convenient and safe, and 4) at substantially lower cost than 1:1 virtual CR visits. Activity promotion apps are effective in various general population applications, causing both the AHA and ESC to call for increased evaluation into the efficacy of app-based physical activity promotion specifically for those referred for CR.^9^ Adapting app content based on clinical review so that it is appropriate and safe for those recovering from TAVR, and then providing content to these patients, is a novel, scalable way to improve timely access to high-quality content to promote physical activity.

Cardio Heart Connect is an 8-week intervention beginning immediately upon referral to CR, designed to be initiated while patients are on waitlists for traditional CR. TAVR recipients engage with Peloton walking, strength, and stretching workouts (not the fitness bike) via a mobile platform to promote physical activity. Peloton was chosen because of its large membership, music libraries, trainer variety, and exercise types, and its availability on iOS and Android platforms. The Cardio Heart Connect platform provides comprehensive CR content and smartwatch integration with a clinically reviewed, pre-set, tailored, and socially engaged set of Peloton modules focusing on walking, strength exercises, yoga breathwork, and stretching, areas of exercise previously used in home-based CR^3^ but not in mobile asynchronous format.

By providing access to a free, tailored version of a widely available fitness application (Peloton) at the time patients are waitlisted for cardiac rehabilitation (CR) following TAVR, and prompting engagement with selected exercise modules over an 8-week period, this study tests a scalable approach to bridging the gap in CR access. We hypothesize that participants randomized to Cardio Heart Connect will demonstrate greater physical activity compared with those on the waitlist receiving attention control, along with improvements in self-reported physical activity, cardiovascular-related quality of life, and cardiovascular health status at 12 months post-randomization. We further anticipate that this approach will facilitate greater subsequent participation in traditional CR, positioning Cardio Heart Connect as a complementary, access-enhancing strategy within the continuum of post-TAVR care.

## Methods

### Conceptual Framework

This study is guided by the CREATE Model of Aging and Technology and the Integrated Theory of mHealth, which together posit that accessible, well-designed mobile health technologies can enhance engagement with health behaviors by improving usability, personalization, and support. Within this framework, a user-friendly, portable platform with strong interface design and technical support facilitates consistent engagement with structured tasks—in this case, prescribed cardiac rehabilitation (CR) exercise modules. Completion of these tasks is expected to increase physical activity, the primary target behavior. Improvements in physical activity are hypothesized to drive downstream gains in key intermediate outcomes, including patient activation, exercise capacity, cardiovascular symptoms, biomarkers, and mental health. These changes ultimately support broader secondary prevention goals, such as increased participation in CR, improved functional capacity and physical fitness, and enhanced quality of life. The framework also recognizes the importance of user characteristics (e.g., age, technology literacy, preferences) and emphasizes the role of high-quality, inclusive, and adaptable design in promoting adoption and engagement across diverse populations.

### Study Design

We will conduct a hybrid type I effectiveness-implementation randomized control trial^11^ with N=200 TAVR patients from the UCHealth Valve Clinic (Figure 1). Using a 1:1 allocation, participants will be randomized to Cardio Heart Connect or the attention control group, designed to control technology access. Cardio Heart Connect is structured to provide Peloton modules and monitoring for 8 weeks. Users are provided an Apple (iOS) or Garmin (Android) smartwatch, based on their personal devices or preferences, for capturing steps (primary physical activity outcome), heart rate, fall detection, etc. At baseline, post-intervention (8 weeks), and follow-up (12 months) we assess participants’ physical activity (daily steps recorded via smartwatch). Aim 1 tests whether Cardio Heart Connect, compared to attention control improves physical activity as measured by daily steps at 8 weeks. Aim 2 examines secondary outcomes relevant to cardiovascular recovery following TAVR: functional capacity (VO2 max; DASI), cardiovascular-related quality of life (KCCQ), and cardiovascular health status (Life’s Essential 8), also at 8 weeks and 12 months. Follow-up time points were chosen to mirror the duration typical of HBCR programs (8 weeks),^6^ to be shorter than typical CR waitlist periods, and align with clinical follow-up. Aim 3 evaluates the implementation of Cardio Heart Connect using a mixed methods approach applying RE-AIM. As such, we will collect and examine survey and interview data from key informants charged with implementing Cardio Heart Connect at the study site throughout the study.

**Figure 1.**
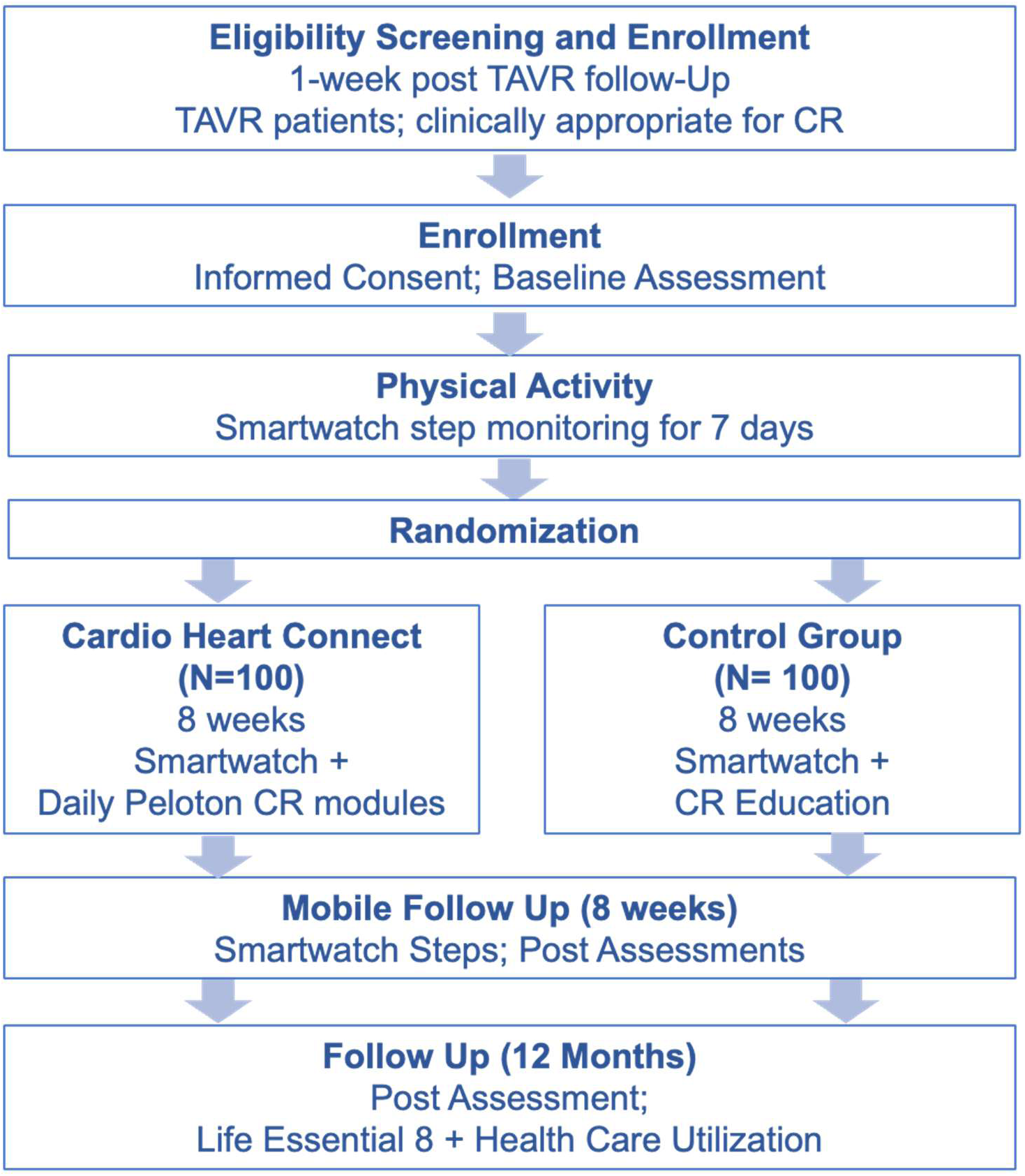
Study Design Overview. Schematic illustrating the overall study design, including participant recruitment, enrollment, data collection, intervention activities, and outcome assessments across the study period.

### Study Setting

The study will take place at the University of Colorado Anschutz Medical Campus. The UCHealth Structural and Valve Clinic participated in several of the landmark trials establishing TAVR as a first-line therapy for severe aortic stenosis and performed the first commercial TAVRs in Colorado.^12, 13^ The clinic continues to perform approximately 200 aortic valve procedures per year.

### Study Population

Participants will include N=200 patients recovering from TAVR from the UCHealth Valve Clinic. Participants will include patients deemed by Valve Clinic staff to be appropriate for CR referral, community-dwelling, live in the United States, and speak English. Participants must be able to provide informed consent and be able to stand with or without an assistive device. Individuals will be excluded for a clinical or self-reported diagnosis of dementia or other higher cognitive impairment, limiting participation. Participants are not excluded due to limited access to technology. We will provide temporary tablets and mobile hotspot connectivity to interested participants who lack hardware or internet access.

### Screening and Recruitment

The UC Structural Heart Clinic will identify potential participants during pre-TAVR evaluation as well as immediate post-procedural recovery and provide them with a study information sheet. Clinical staff will The Structural Heart Program Advanced Practice Nurse will conduct a screening of all TAVR patients who are scheduled for their pre-TAVR visit and assess preliminary eligibility via the electronic health record (EHR). Based on our previous work recruiting for similar trials, we estimate 90% of patients recovering from TAVR will be eligible, and 43% will enroll, as many as 77 patients per year, indicating our capacity to enroll N=200 patients in 3 years to allow for 12-month follow-up. During the patient’s pre-TAVR visit, research staff will meet with the potential participants, explain the study, and invite them to participate.

### Enrollment and Baseline Visit

Based on the patient’s preference and availability, participants can complete their baseline visit in person during TAVR discharge, at the post-TAVR 1-week appointment, or schedule a Zoom enrollment visit up to 1-month following their TAVR. During the enrollment/baseline visit, participants will complete an electronic informed consent in REDCap, via a tablet if in-person or via Zoom. The research assistant will help participants create user accounts to access study materials and smart watch data, download apps, and demonstrate features/tools on phone, tablet, and/or computer. Baseline assessment (Table 2) will be collected via REDCap, and a smartwatch will be provided/mailed to wear for seven consecutive days. Participants are required to wear the watch for 7 days, a minimum of 10 hours for 4 of the 7 days, to accurately assess daily steps.

**Table 1.**
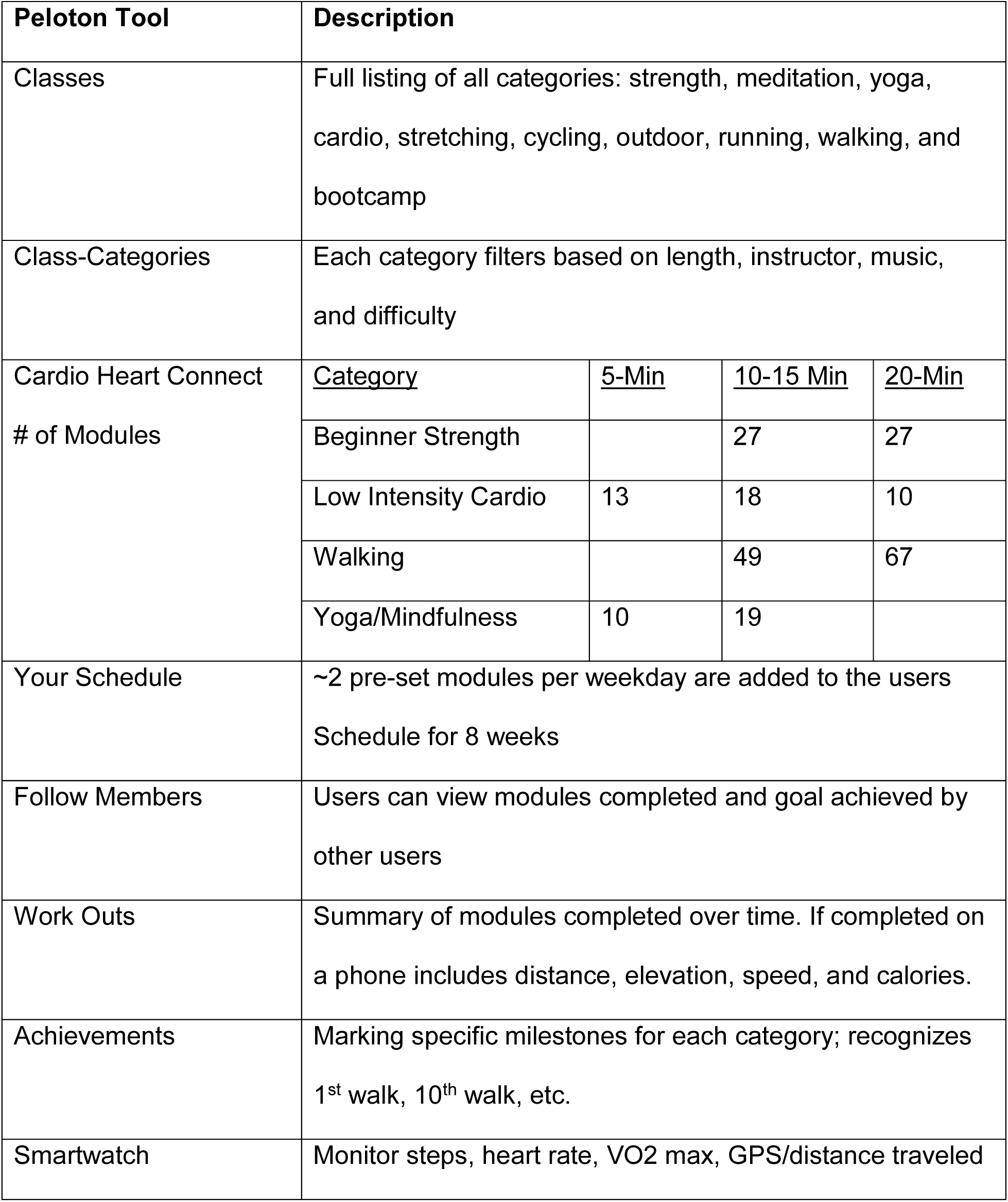
Summary of Cardio Heart Connect Tasks.

**Table 2.** Control Weekly Schedule of ACC informational resources.

|  |  |
| --- | --- |
| Week 1 | Discuss cardiac diagnosis – heart and valve disease |
| Week 2 | Cholesterol management |
| Week 3 | Physical activity/ “move more” |
| Week 4 | Nutrition and Diet |
| Week 5 | Medication compliance |
| Week 6 | Healthy lifestyle – smoking cessation and alcohol intake |
| Week 7 | Healthy lifestyle – stress management, sleeping habits, and mindfulness/managing energy |
| Week 8 | Post-program planning – cardiac rehab and community exercise programs / “Action Plan” |

**Table 3.**
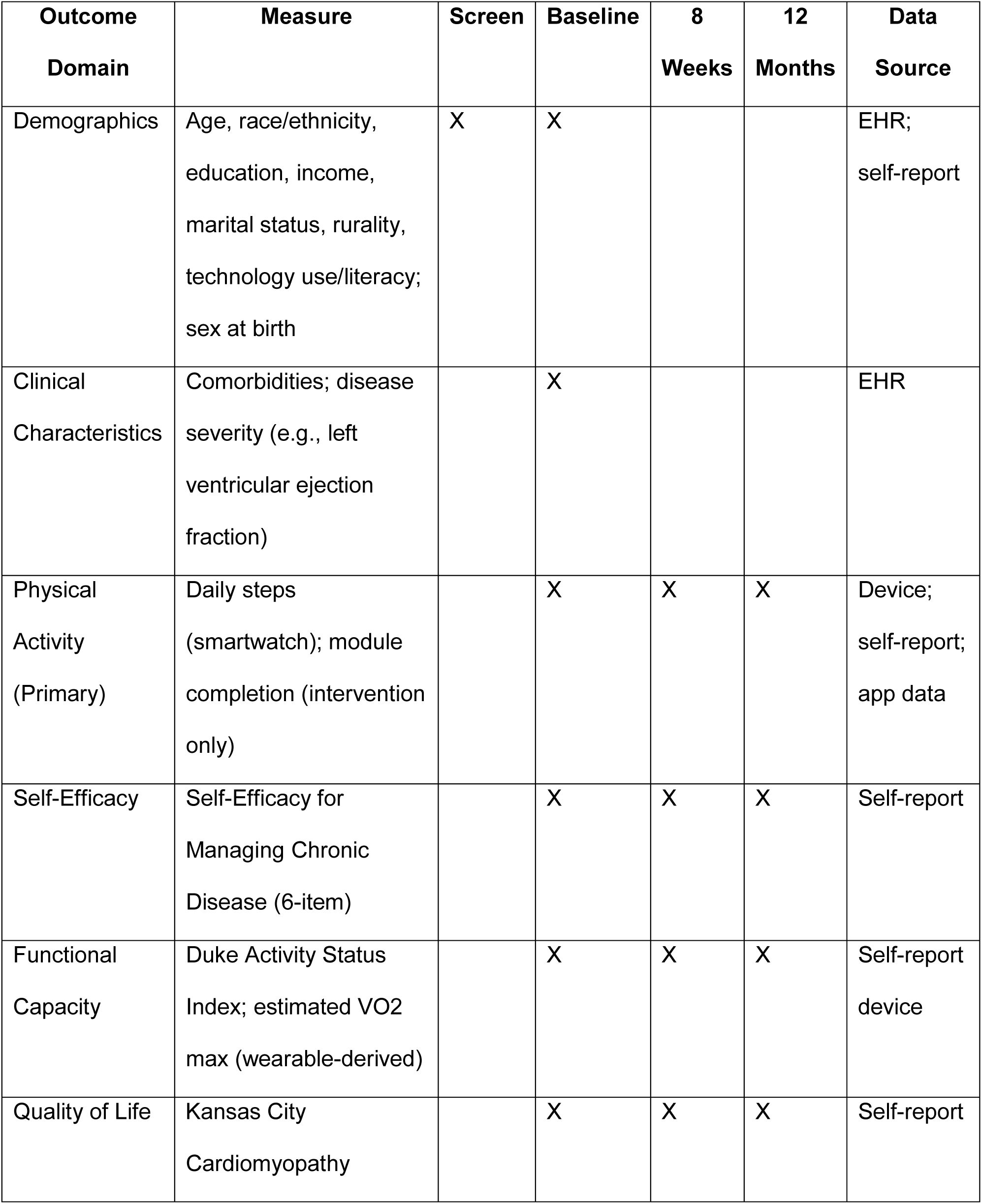
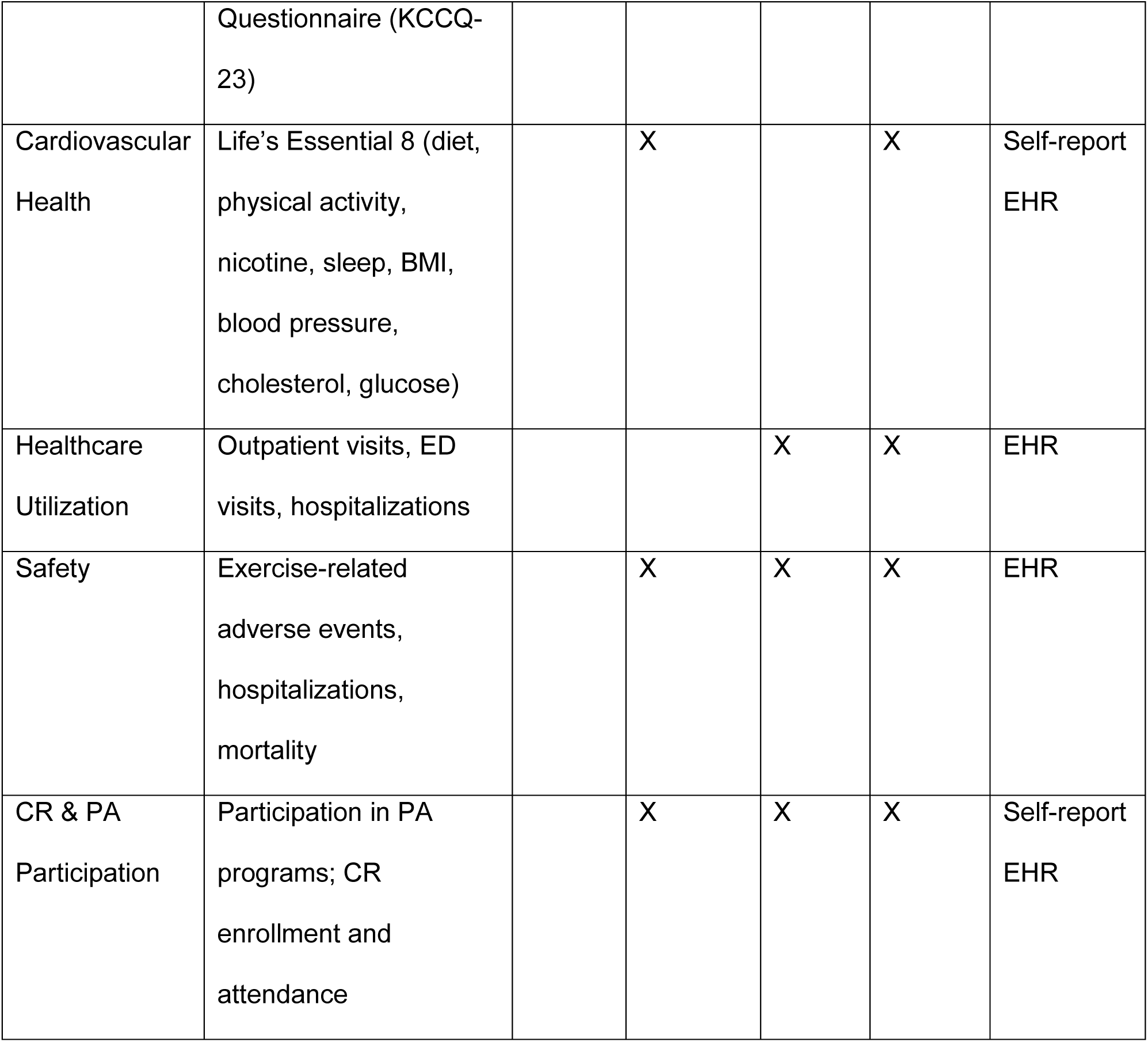
Measures and Assessment Timeline.

### Randomization and Blinding

After the 7-day wear period, the research assistant(s) will meet with participants via Zoom to verify baseline assessment completion and step capture via the smartwatch. The participants are then randomly assigned using a 1:1 allocation to Cardio Heart Connect or control group. Research assistants will then provide a tutorial for using either Cardio Heart Connect or inquire about the control participant’s experience with the watch. Although study participants and team members executing the recruitment are not blinded, outcome assessors and the principal investigators are blinded to the intervention allocation. We instruct study participants not to discuss their study allocation with those team members assessing the outcomes.

### Intervention: Cardio Heart Connect

Participants will engage with CR-related Peloton modules - ranging in duration from 5 minutes in early phases to 20 minutes in later phases - for 8 weeks. Each patient will be scheduled ∼80 modules focusing on walking, strength, yoga (meditation and breathwork), and stretching, areas of exercise training previously found effective in HBCR and individualized by the research team based on patient preferences (e.g. music, coaches, more or less yoga content). The modules were systematically reviewed by the research team and curated to ensure they are appropriate for TAVR recovery.

Move in a Tube Pathway: Some estimates hold that as many as a third of patients receiving TAVR require new pacemaker placement following their procedure^14^. These patients will still be eligible for the trial, and therefore common activity restrictions in the post-implantation period necessitate the creation of another treatment pathway. The “Move in a Tube” approach allows patients to participate in similar levels of activity guided by Peloton modules, but which are restricted further to avoid movements which require patients to lift their arms above their shoulders (e.g. shoulder presses, yoga) or extending outward from their torso (e.g. push ups). These guidelines ensure patient safety while simultaneously promoting physical rehabilitation. Research staff created a separate set of Peloton workout exercises for patients to complete that adhere to Move in a Tube Pathway protocols as confirmed by clinicians. Once a patient is randomized into the intervention group, if the patient’s discharge summary includes a pacemaker notation, they are enrolled in the “Move in a Tube” Peloton pathway. If they do not have a pacemaker, they are enrolled in the conventional Peloton pathway.

For each weekday, participants will be prompted via text to complete 2 Peloton modules totaling 10-25 minutes per day and progressing over difficulty time. They can also review “Your Schedule” to re-complete modules based on preferences. We will encourage participants to only use the recommended modules; however, it is possible they will try additional modules.

Since the study team creates and controls the Peloton accounts, we have full access to modules completed and notification, sharing, and support functions. Participants will be semi-supervised by the research team as heart rate and falls are monitored daily by the team and follow up as needed. Participants are prompted to report any adverse event(s) to research staff, which are used for reporting to the DSMB and to improve the experience for future trials. Participants are required to wear the smartwatch during each module, to monitor fall detection during the session. Research assistant(s) review module completion, feedback, heart rate information, and fall detection indicators. If any falls are detected, the research team contacts the participant by phone to confirm participant safety and follow emergency support and reporting protocols.

### Control Group: Cardio Heart Education

Control participants have access to a smartwatch and a web-based platform that provides weekly emails sharing CR education materials. The weekly Cardio Heart Education program messages the participant with resources available from the American College of Cardiology’s patient-facing CardioSmart website. Education content was chosen specifically to address cardiovascular health and CR following transcatheter procedures. Table 2 specifies the schedule of when these resources will be made available to the Control group.

### Retention Strategies

We encourage sustained research participation and adherence using strategies previously shown to be beneficial in mHealth trials,^15,16^ including: (a) deploying customizations and refinements to data capture and intervention platforms that directly responds to user feedback; (b) stressing the significance of participation as a contribution to future patients’ wellbeing; (c) providing a library of detailed instructions and demonstration on how to use all parts of the system including Peloton and platform interfaces; (d) providing readily accessible technology-support through a tech-support hotline; (e) using a participant tracking database to prompt participation to complete study tasks; (f) issuing study appointment (baseline, post-intervention, and 12 months) reminders via text, email, and telephone; and (g) providing modest monetary incentives for follow-up.

### Study Assessments and Procedures

Our selected outcomes are based on earlier trials of home-based cardiac rehabilitation programs^7^ and our conceptual framework.

#### Primary Outcome

##### Accelerometer Daily Steps

Average daily steps will be measured objectively over the 7-day periods using the Apple and Garmin smart watches, which have both been validated as highly accurate activity monitors, with high inter-device reliability for step calculation.^17^ The watches quantify free-living sedentary and ambulatory activities to estimate time spent sitting/lying, standing, and stepping. Participants will wear the device for seven continuous days immediately following each study visit. Days with at least 10 hours of wear will be considered valid, and participants must have a minimum of 4 valid days (including 1 weekend day) for data to be included. The time-stamped “daily step-count” data will be used, all averaged over the 7 days. Additional digital measures for physical activity and functional capacity, including VO2max, will be captured from the smartwatches; however, these measures are not as well validated as steps but will be available for secondary analyses.^17, 18^

*Duke Activity Survey Index (DASI) is* a 12-item self-report questionnaire measuring functional capacity including personal care, ambulation, household tasks, sexual function, and recreation. Criterion validity data describe significant correlations with peak oxygen intake (*r = .80)*, health related quality of life (*r = -.64)*, and depressive symptoms (*r = -.44)* in patients with cardiovascular disease. Internal consistency reliability is high (α=.86). Each item is weight-based on “yes” responses and summed for a total score. Higher scores indicate better functional status. Total scores range between 0 - 58.2. The DASI matched metabolic expenditure (METS) scores range *between 0 - 9.89.*^14, 19^

##### Kansas City Cardiomyopathy Questionnaire (KCCQ)

The KCCQ is a 23-item scale developed to aid in rapid assessment of patient-relevant QOL facets in CV care. Validity data were initially described through convergent validity with existing longer measures of health status. Reliability statistics for each subscale all sit above .6, including those for physical limitation 46 (α=.90), symptoms (α=.88), QOL (α=.78), social limitation (α=.86), and self-efficacy (α=.62). Most critical to this project, reliability for summary scores sits considerably higher, including for functional status (α=.93), and overall KCCQ summary score (α=.95). Each scale score ranges from 0-100 with increased scores denoting improved quality of life and general level of functioning.^20–22^

##### Life Essential 8

Defined by the American Heart Association, the Life’s Essential 8 is an 18-item measure outlining health behaviors (healthy diet, physical activity, nicotine exposure, and sleep) and health factors (body mass index, blood pressure, total cholesterol, and blood fasting glucose) that are associated with cardiovascular health.^23,24^ Health behaviors are captured via self-report. Physical activity, PAQ-K, and sleep are measured as described above. Healthy diet is captured by the DASH (Dietary Approaches to Stop Hypertension trial) Diet Score^25^, nicotine exposure by the National Health and Nutrition Examination Survey - smoking and tobacco use (NHANES SMQ)^26^, physical activity by the NHANES Physical Activity Questionnaire (PAQ), and sleep by the NHANES Sleep Disorder Questionnaire (SLQ). Health factors are collected as part of clinical care and extracted from the EMR at baseline and patient’s 12-month TAVR follow up. Each domain has an algorithm based on healthy thresholds ranging from 0 to 100 points. A composite score varies from 0 to 100 with higher scores indicating better cardiovascular health status.

### Data Analysis

#### Power Analysis and Sample Size

The power analysis was conducted using the Aim 1 physical activity outcome of daily steps at 8 weeks and 12 months. Studies using commercial wearable monitors indicate very good participant retention; for example, a large study of older adults found 92% of users to have sufficient data for analysis (at least 4 days of valid wear in a 7-day period). Our proposed sample size of 200 is sufficient to detect a small to moderate effect size (Cohen’s D =0.4) with approximately 80% power while allowing for 8% attrition. Assuming a standard deviation of 2,500 steps per day,^27,28^ this effect size corresponds to a difference of 1,000 daily steps between the control and intervention groups, which is a clinically meaningful difference. For secondary analyses of physical activity maintenance at 12 months, we anticipate a lower sample size of around 150; power to detect a difference of 1,000 daily steps will therefore decrease to 0.65, but we will be able to detect a difference of 1,200 daily steps between groups (Cohen’s D = 0.48) with 80% power. Power calculations were conducted using R version 4 (R Core Team, 2020).

Aim 2 analyses are exploratory. For the KCCQ, assuming a standard deviation of 20 in this population, we have sufficient power to detect a difference of 8 points on a 100-point scale; for the DASI, assuming a standard deviation of 15, we have sufficient power to detect a difference of 6 points (e.g., from 27 to 33 on the 0 to 58.2 point scale). The use of VO2max as a wearable-assessed outcome is far more nascent, but previous studies of HBCR programs have illustrated improvements of 1.2mL/kg at 6 months attributable to remote activity promotion.^7^ The Life’s Essential 8 was released by the AHA in June 2022, so current knowledge of the distribution and clinically important effect sizes are limited. However, one initial study suggests that the Essential 8 is more sensitive to differences between groups than the previous Life’s Simple 7, for which differences as low as 1 point on a 14-point scale were associated with significant differences in cardiovascular outcomes. For Aim 3, we anticipate that the sample size computed for Aim 1 will allow for sufficient assessment of implementation.

#### Data Preparation and Missing Data Analysis

Efforts to minimize missing data include incentivizing participation and research assistant review during baseline and follow-up visits. The project manager assesses participant engagement throughout the 8-week study period and assists with any technical issues. The project analyst performs data quality checks every 6 months during periods of active data collection and includes checks for missing data fields. Prior to data analyses, data will be examined to consider whether missing data are ignorable (missing completely at random (MCAR) or missing at random) or are non-ignorable (missing not at random (MNAR)) using Little’s test of MCAR and sensitivity analyses using varying MNAR mechanisms. We anticipate that missing data will be MCAR or MAR; the proposed repeated mixed effects model produces unbiased parameter estimates with these missing data mechanisms.

#### Analysis Plan

Descriptive statistics will include means and standard deviations for normally-distributed continuous variables, medians and interquartile ranges for continuous variables that are not normally distributed, and frequencies and percentages for categorical variables. These will be presented both overall and by treatment group; we will compare demographics and baseline predictors between treatment groups using independent samples t-tests, Wilcoxon-Mann-Whitney tests, and Chi-square tests as appropriate, to identify any covariates which are unbalanced between treatment groups for inclusion in our mixed models.

The analyses of effectiveness will use repeated measures mixed models. Each analysis model will include an indicator variable for the treatment group, a random effect for subject, and will control for baseline covariates (if any) which are found to unbalanced between the treatment and control groups. Repeated measures mixed models allow for partially incomplete data (e.g., a subject missing follow-up for one of the collection periods) and relaxes the missing data assumptions to missing at random conditional on observed data. Primary analyses will compare outcomes at baseline and 8 weeks only; secondary analyses will include the 1-year timepoint to assess maintenance.

All statistical analyses will be conducted using R (R Core Team, 2020) or SAS v9.4 (SAS Software, Cary, NC, USA). A significance level of 0.05 will be used for all tests and confidence interval estimates.

### RE-AIM Evaluation

Implementation and evaluation of the intervention are guided by the RE-AIM Framework, which examines Reach, Effectiveness, Adoption, Implementation, and Maintenance in the context of multilevel influences, including external factors, health system infrastructure, and patient and clinician characteristics (Table 4). RE-AIM will be applied across the planning, implementation, and evaluation phases using a mixed methods approach. Quantitative data will assess participation, outcomes, and fidelity, while qualitative interviews with patients, clinicians, and health system leaders will provide contextual insights into barriers, facilitators, and real-world applicability. Rapid qualitative analysis will be used to efficiently synthesize findings and inform iterative refinements to study procedures, recruitment strategies, and technical support throughout the trial.

**Table 4.** Summary of RE-AIM Evaluation of Cardio Heart Connect (CHC)

| Domain | Data Source | Quantitative Measure / Example Questions |
| --- | --- | --- |
| Reach | Recruitment<br>Database | # screened, eligible, enrolled; reasons for non-enrollment |
| Effectiveness | Aim 1 & Aim 2 | Efficacy testing of CHC on health outcomes |
| Adoption | Partner Interviews<br>(Clinicians) | • What are best practices for clinical referral to CHC? Are there reasons not to refer?<br>• Are resources (personnel, system-level, technical) in place for CHC?<br>• What patient and organizational barriers were there? |
| Implementation | App use data;<br>Fidelity<br>Acceptability<br>Partner Interviews<br>(Clinicians & Patients);<br>Adaptations;<br>System costs | # log-ins, features used, adherence to CHC<br>Fidelity to intervention delivery, content, technical support<br>Descriptive analysis of uMARS<br>• Describe your experience with CHC; how was it used/modified?<br>• What costs or resource needs should be considered? |
| Maintenance | Aim 2; Partner Interviews<br>(Clinicians & Health System Leaders) | Exploratory efficacy testing of CHC at 12 months<br>• How would CHC fit within workflows?<br>• How could CHC be sustained or scaled? |

Reach will be evaluated by the proportion and representativeness of eligible patients who participate, with attention to equity across key demographic and clinical characteristics. Effectiveness will be assessed through primary and secondary outcomes described in the trial aims. Adoption will focus on clinician engagement with referral processes and reasons for non-participation. Implementation will include evaluation of fidelity, adaptations, costs, and acceptability, incorporating both quantitative metrics (e.g., adherence, engagement, staff effort) and participant-reported usability. Maintenance will be examined at both the patient level (12-month outcomes) and system level, including stakeholder perspectives on sustainability and scalability. Together, these data will inform the feasibility, generalizability, and long-term integration of the intervention into routine care.

### Ethical and Safety Considerations

All study procedures involving human participants will be conducted in accordance with established ethical standards for research. The protocol has been reviewed and approved by the Colorado Multiple Institutional Review Board (COMIRB; Protocol #24-0626), which will provide ongoing oversight and monitoring throughout the study. Informed consent will be obtained from all participants prior to enrollment, and appropriate safeguards will be in place to protect participant privacy, confidentiality, and data security.

#### Potential Risks

Potential adverse events include injuries sustained while exercising, including falling, tripping or other injuries during Peloton modules, which may occur as participants engage with walking, strength, and stretching modules. Psychological discomfort may arise during self-report assessments of quality of life and mental health measures, and participants who screen positive for depressive symptoms (KCCQ-23 items 12 and 14 score <= 4) are provided a recommendation to discuss emotional concerns with their health care provider. Increased pain or other symptoms from increased physical activity may occur as participants begin the intervention, potentially experiencing muscle or joint discomfort; delayed onset muscle soreness (DOMS) is particularly common when people begin a new exercise program, especially if they are relatively sedentary before the intervention, which may be perceived as increased pain or related symptoms and could lead to fear of worsening physical symptoms and subsequent risk of dropping out of the intervention. Loss of confidentiality is also a potential risk, as study team members have access to Peloton log-in information and participants are using a platform with social and sharing features that could potentially expose their workout activity to others.

#### Adverse Event Reporting

Each adverse event (AE), serious adverse event (SAE), and Unanticipated Problem (UP) are reported according to standard NHLBI and COMIRB procedures, including the study team’s assessment of their severity and plausible causal connection to study participation. Research subjects involved in this proposal are given contact information for study staff should they experience any potential adverse side effects or concerns.

#### Defining and Reporting of Serious Adverse Events (SAEs)

We follow local guidelines that require investigators to promptly notify the IRB (within 5 days of the initial receipt of information) when SAEs occur. SAEs are defined as death, life threatening illness, hospitalization, or any event which results in the individual being unable to performing usual activities of daily living (ADLs). These are distinct from AEs which do not result in any participant activity limitations (deemed “mild”), or that interfere with, rather than prevent, ADL’s (deemed “moderate”). The IRB requires that any SAE that is unexpected and possibly related to a research intervention must be reported. SAEs that are unrelated to trial participation will not be reported. Risks that are described in the protocol and consent form do not have to be reported as SAEs, unless the expected SAE occurs more frequently or is more serious than expected. One exception to this rule is in the case of death; all deaths must be reported.

#### Discontinuation and Participant Withdrawal

DSMB members are immediately notified of any study-related SAEs, and review AEs at regular quarterly meetings. In conjunction with the DSMB, the investigators will discontinue or stop the study if there is deemed to be a legitimate concern about ongoing safety risks to trial participants. The DSMB has access to outcome data. Because there are no known adverse effects associated with the planned intervention, no interim analysis of the data is planned. The DSMB has the right to consider interim analyses as requested. These procedures could include guidelines for early termination for benefit, termination for futility, and termination for safety reasons.

## Discussion

Physical activity is a critical piece of recovery from TAVR, yet traditional cardiac rehabilitation is often inaccessible and/or doesn’t meet patients at the right time to catch them in a moment that is likely to maximize their benefit. Asynchronous exercise has illustrated feasibility in cardiovascular risk prevention^29^ in part due to its flexibility; it does not have to be conducted during work hours or in medical centers, unlike traditional CR.

It is imperative to make efforts to reduce the disparity in CR engagement, especially among those in medically underserved communities. Even moderate increases in physical activity are likely to reap benefits in terms of patient outcomes. Cardio Heart Connect uses commercially-available fitness content and off-the-shelf technology that is accessible and usable among most patients. Trial results will help provide evidence for and justify further expansion of technological access for patients.

### Limitations

Our study has potential limitations. Recruitment for the Cardio Heart Connect study is being held out of an academic medical center, and thus the experience and results might not be generalized to those cared for in other settings. Additionally, patients receiving TAVR, who are then referred to CR, are likely marginally healthier and more physically capable at the outset than many patients at initial referral to CR. Care must be exercised when interpreting any findings from Cardio Heart Connect to populations with different CR indications.

## Conclusions

Despite ample evidence of the benefits of CR, only a small fraction of TAVR patients participate. This study tests the use of clinically controlled, commercially available fitness programming, Cardio Heart Connect that targets physical activity and cardiovascular health outcomes. Commercially available mobile health provides an opportunity to deliver physical activity promotion content to patients challenged to participate in traditional CR.

## Data Availability

Data generated from this study will be made publicly available in accordance with NIH Data Management and Sharing Policy requirements following completion of the trial and publication of the primary study results. De-identified participant-level data and accompanying documentation will be deposited in an appropriate data repository and made available to qualified researchers consistent with applicable regulatory, ethical, and privacy considerations.

## Acknowledgements

We would like to thank all the patients who have dedicated their time to this study. The authors would also like to thank the clinicians at the UCHealth Heart and Vascular clinic, as well as the clinicians involved in this study. The authors would like to acknowledge Geoffrey Harger, MPH, for his contributions to project management and trial preparation.

## Sources of Funding

This study is supported by the National Heart, Lung, and Blood Institute (R61HL169336/R33HL169336). Dr. Knoepke was also supported under a separate award from National Heart, Lung, and Blood Institute (K23HL153892) during the course of this project. The project discussed here, and all opinions are solely those of the investigators and not of any funding agency, including the NIH. The funding bodies had no role in the design of the study, analyses, interpretation of data, drafting or editing of manuscripts, or decisions to submit for publication.

## Disclosures

None.

## Notes

### Competing Interest Statement

The authors have declared no competing interest.

### Clinical Trial

NCT07008911

### Author Declarations

The protocol has been reviewed and approved by the Colorado Multiple Institutional Review Board (COMIRB; Protocol #24-0626), which will provide ongoing oversight and monitoring throughout the study. Informed consent will be obtained from all participants prior to enrollment, and appropriate safeguards will be in place to protect participant privacy, confidentiality, and data security.

